# Longitudinal changes in home-based arts engagement during and following the first national lockdown due to the COVID-19 pandemic in the United Kingdom

**DOI:** 10.1101/2021.05.14.21257233

**Authors:** Feifei Bu, Hei Wan Mak, Jessica K Bone, Daisy Fancourt

**Affiliations:** Department of Behavioural Science and Health, University College London, UK

**Author notes:** Corresponding author: Daisy Fancourt, Address: 1-19 Torrington Place, London, UK, WC1E 7HB.

**Keywords:** health promotion, arts engagement, COVID-19, lockdown, longitudinal, growth trajectory

## Abstract

**Aims:** This study aimed to examine potential heterogeneity in longitudinal changes in home-based arts engagement during the first national lockdown and following gradual easing of restrictions in the UK. Further, it sought to explore factors that were associated with patterns of longitudinal changes in home-based arts engagement.

**Methods:** Data were from the UCL COVID-19 Social Study. The analytical sample consisted of 29,147 adults in the UK who were followed up for 22 weeks from 21^st^ March to 21^st^ August 2020. Data were analysed using growth mixture models.

**Results:** Our analyses identified five classes of growth trajectories. There were two stable classes showing little change in arts engagement over time (64.4% in total), two classes showing initial increases in arts engagement followed by declines as restrictions were eased (29.8%), and one class showing slight declines during strict lockdown followed by an increase in arts engagement after the easing of restrictions (5.9%). A range of factors were found to be associated with class membership of these arts engagement trajectories, such as age, gender, education, income, employment status, and health.

**Conclusions:** There is substantial heterogeneity in longitudinal changes in home-based arts engagement. For participants whose engagement changed over time, growth trajectories of arts engagement were related to changes in lockdown measures. These findings suggest that some individuals may have drawn on the arts when they needed them the most, such as during the strict lockdown period, even if they usually had lower levels of arts engagement before the pandemic. Overall, our results indicate the importance of promoting arts engagement during pandemics and periods of lockdown as part of public health campaigns.

## Introduction

The outbreak of COVID-19 pandemic had a substantial impact on the cultural sector. The introduction of lockdown and “stay-at-home” orders led to the closure of public spaces, galleries, museums, arts venues, and other cultural assets. However, the pandemic also provided new opportunities to engage in the arts at home through both increased digital availability of the arts (e.g., virtual choirs and online arts classes) and the introduction of furlough schemes, whereby large proportions of the population were required to take leave from work. Home-based arts engagement may therefore have increased during the pandemic. In the first United Kingdom (UK) lockdown, the online sales of a large arts and crafts retailer increased by 200% [1]. Additionally, 22% of people reported spending more time on home-based arts activities during the first UK lockdown, and 52% of these people maintained or increased these levels of arts engagement three months later [2].

Given the well-established association between arts engagement and health, changing patterns of arts engagement may have important implications for health and wellbeing. Arts engagement involves health-promoting activities that may, in turn, activate a number of psychological processes (e.g. improving self-esteem), physiological processes (e.g. reducing stress hormones), social processes (e.g. reducing loneliness), and behavioural processes (e.g. enhancing agency; [3]). Through these mechanisms, arts engagement may contribute to the promotion of health and wellbeing, prevention of mental and physical illness, and management of existing health conditions [4]. There is evidence that the arts have played an important role in supporting wellbeing specifically during the COVID-19 pandemic. For example, engagement in hobbies (e.g. reading and listening to the radio/music) was associated with lower levels of depressive and anxiety symptoms and higher levels of life satisfaction in adults in the UK [5]. Similarly, musical activities helped improve people’s mood, boosted their confidence and sense of positivity, and led to a greater sense of connectedness in adults in Spain [6]. It has been proposed that people used the arts to help cope with their emotions and improve their self-development during lockdown [2].

Whilst there appears to have been an overall increase in arts engagement during initial COVID-19 lockdowns, engagement may have been socially patterned [2, 7]. Pre-pandemic studies have repeatedly found that arts engagement is higher amongst younger adults, women, people living in rural areas, those with higher educational levels, and individuals with greater social support [8–11]. Many of these groups have also made greatest use of the arts during the COVID-19 pandemic [2]. However, there is some evidence that other factors such as ethnicity, partnership status, socio-economic status, and mental/physical health conditions were differentially associated with arts engagement prior to and during the COVID-19 pandemic [2]. For example, ethnicity was not associated with arts engagement during the first UK lockdown [2], despite previous evidence that people from an ethnic minority background engaged in the arts less prior to the pandemic [9, 10]. Also in contrast to previous findings, people with higher levels of loneliness and diagnosed mental health conditions had higher engagement levels [2]. This suggests that new profiles of arts audiences might have emerged during the pandemic.

However, several questions remain unanswered. To date, research has focused on average levels of arts engagement during COVID-19, conflating the nuanced experiences of different subgroups and how these experiences might have evolved longitudinally. It is also unclear whether the different stages of lockdown, such as the easing of restrictions, led to changes in arts engagement. Understanding the longitudinal patterns of arts engagement during the pandemic, and individual and societal factors associated with these patterns, is crucial for understanding how and when individuals use the arts to support their wellbeing. It is also important for the arts sector to understand how the pandemic affected patterns of arts engagement. Identifying whether any changes in engagement were temporary, whilst social restrictions were most stringent, or have persisted following the easing of restrictions may show whether audiences for home-based arts activities have changed. This could guide strategies for arts funding and broader cultural policies to re-establish the cultural sector as the pandemic continues and abates [12].

In light of this, the present study aimed to examine how home-based arts engagement changed during the COVID-19 pandemic in the UK. First, we investigated potential heterogeneity in longitudinal changes in arts engagement, using a large sample of 29,147 adults followed across 22 weeks from 21^st^ March to 21^st^ August 2020, a period that spanned whole of the first lockdown and the easing of restrictions. Second, we explored whether a range of factors were associated with different patterns of longitudinal changes in arts engagement.

## Methods

### Sample

We analysed data from the UK COVID-19 Social Study run by University College London; a longitudinal study that focuses on the psychological and social experiences of adults living in the UK during the COVID-19 pandemic. The study commenced on 21st March 2020 and involves weekly and then monthly online data collection from participants for the duration of the pandemic. The study did not use a random sample design and therefore the original sample is not representative of the UK population. However, it does contain a heterogeneous sample that was recruited using three primary approaches. First, convenience sampling was used, including promoting the study through existing networks and mailing lists (including large databases of adults who had previously consented to be involved in health research across the UK), print and digital media coverage, and social media. Second, more targeted recruitment was undertaken focusing on (i) individuals from a low-income background, (ii) individuals with no or few educational qualifications, and (iii) individuals who were unemployed. Third, the study was promoted via partnerships with third sector organisations to vulnerable groups, including adults with mental health conditions, older adults, carers, and people experiencing domestic violence or abuse. A full protocol for the study is available online at www.COVIDSocialStudy.org.

We included participants who had at least three repeated measures between 21^st^ March and 21^st^ August 2020 when the study switched from weekly to monthly follow-up and the relevant measure was discontinued (49,846 participants). Around 10% of these participants withheld data or preferred not to report on demographic and health-related factors and were therefore excluded from our analysis. Further, we excluded participants (32%) with missing data on comparative arts engagement (during vs pre-pandemic). This provided us a final analytic sample size of 29,147 participants, followed-up for a maximum of 22 weeks. See the Supplementary Material for an overview of the UK COVID-19 restrictions during this period.

### Measures

Arts engagement was measured by asking how long participants had spent engaging in a home-based arts or crafts activity (e.g., painting, creative writing, sewing, playing music, etc) on the last working weekday. Asking about the last working weekday aimed to encourage specificity of recall, following the ‘time diary’ approach, and remove variation from those who took part on weekends [13]. Weekly responses were recorded on a five-point frequency scale, from did not do to ≥6 hours. Given the low frequency of arts engagement, we created a binary variable indicating whether participants spent any time on arts or crafts activities during the last working day (yes vs no).

A range of socio-demographic and health-related factors measured at baseline were considered as predictors of arts engagement trajectories. These included gender (women vs men), ethnicity (white vs ethnic minorities), age (18-29, 30-45, 46-59, 60+ years), education (GCSE levels or below, A-levels or equivalent, degree or above), household income (<£30,000 vs >£30,000 per annum), employment status (employed throughout, employed at baseline but lost job during the follow-up, unemployed or economically inactive), living arrangement (living alone, living with others but no children, living with others including children), area of living (city, large town, small town, rural). Health-related factors were self-reported disability (yes vs no) and self-reported diagnosis of a long-term mental health condition (yes vs no). We also included a comparative measure of arts engagement, in which participants were asked how their current arts engagement (measured 21^st^-27^th^ May 2020) compared to their engagement levels before the pandemic (less than usual, about the same, more than usual).

### Statistical analysis

Data were analysed using the growth mixture modelling (GMM) approach. The conventional growth modelling approach specifies one homogeneous growth trajectory, allowing individual growth factors to vary randomly around the overall mean. GMM relaxes this assumption and enables researchers to explore different patterns of longitudinal changes (latent trajectory classes; [14]).

Starting with the unconditional GMM, we compared models with different numbers of classes using the Bayesian information criterion (BIC), sample-size adjusted Bayesian information criterion (ABIC), Vuong-Lo-Mendell-Rubin likelihood ratio (LMR-LR) test, and adjusted Lo-Mendell-Rubin likelihood ratio (ALMR-LR) test. We included quadratic and cubic functions of time scores to allow for nonlinear polynomial changes. After identifying the optimal number of classes, we introduced predictors to explain the observed heterogeneity between classes.

Weights were applied throughout the analyses. The final analytical sample were weighted to the proportions of gender, age, ethnicity, education and country of living obtained from the Office for National Statistics [15]. The descriptive analyses were implemented in Stata v16 and GMM in Mplus Version 8.

## Results

Before weighting the 29,147 participants, there was an over-representation of women and people with a degree or above and under-representation of people from ethinic minority backgrounds and adults under 30 (Table 1). After weighting, the sample reflected population proportions, with 50.6% women, 33.4% with a degree or above, 12.8% of ethnic minority and 19.5% under 30. Figure 1 shows the percentage of participants who spent time on arts activities over the 22-week follow-up.

**Table 1.** Descriptive statistics of the sample (N=29,147)

|  | Raw data |  | Weighted data |  |
| --- | --- | --- | --- | --- |
|  | % | N | % | N |
| **Gender** |  |  |  |  |
| Women | 74.6% | 21,735 | 50.6% | 14749 |
| Men | 25.4% | 7,412 | 49.4% | 14398 |
| **Ethnicity** |  |  |  |  |
| Minority | 3.9% | 1,136 | 12.8% | 3731 |
| White | 96.1% | 28,011 | 87.2% | 25416 |
| **Age** |  |  |  |  |
| 18-29 | 5.6% | 1,635 | 19.5% | 5684 |
| 30-45 | 24.2% | 7,052 | 26.1% | 7607 |
| 46-59 | 32.4% | 9,454 | 24.1% | 7024 |
| 60+ | 37.8% | 11,006 | 30.3% | 8832 |
| **Education** |  |  |  |  |
| Low (GCSEs or below) | 12.9% | 3,774 | 32.7% | 9531 |
| Medium (A-levels or equivalent) | 16.7% | 4,868 | 33.9% | 9880 |
| High (Degree or above) | 70.4% | 20,505 | 33.4% | 9736 |
| **Household income (annual)** |  |  |  |  |
| <16k | 14.4% | 4,198 | 19.2% | 5587 |
| 16-29k | 25.1% | 7,309 | 27.8% | 8105 |
| 30-59k | 35.3% | 10,294 | 33.0% | 9619 |
| 60-89k | 14.9% | 4,342 | 12.2% | 3566 |
| ≥90k | 10.3% | 3,004 | 7.8% | 2270 |
| **Employment status** |  |  |  |  |
| Employed | 50.5% | 14,728 | 46.6% | 13594 |
| Employed to unemployed | 10.1% | 2,955 | 10.6% | 3076 |
| Unemployed/economically inactive | 39.3% | 11,464 | 42.8% | 12477 |
| **Living status** |  |  |  |  |
| Living alone | 21.8% | 6,366 | 19.0% | 5550 |
| Living with others (not children) | 55.5% | 16,171 | 57.3% | 16707 |
| Living with others (including children) | 22.7% | 6,610 | 23.6% | 6890 |
| **Area of living** |  |  |  |  |
| City | 32.8% | 9,571 | 34.3% | 10011 |
| Large town | 17.5% | 5,096 | 20.6% | 6011 |
| Small town | 25.3% | 7,373 | 24.9% | 7257 |
| Rural area | 24.4% | 7,107 | 20.1% | 5868 |
| **Disability** |  |  |  |  |
| Yes | 7.8% | 2,279 | 9.4% | 2748 |
| No | 92.2% | 26,868 | 90.6% | 26399 |
| **Mental health diagnosis** |  |  |  |  |
| Yes | 16.8% | 4,907 | 20.1% | 5856 |
| No | 83.2% | 24,240 | 79.9% | 23291 |
| **Arts engagement** |  |  |  |  |
| Less than usual | 17.8% | 5,197 | 15.9% | 4641 |
| About the same | 56.7% | 16,524 | 61.2% | 17851 |
| More than usual | 25.5% | 7,426 | 22.8% | 6655 |

**Figure 1.**
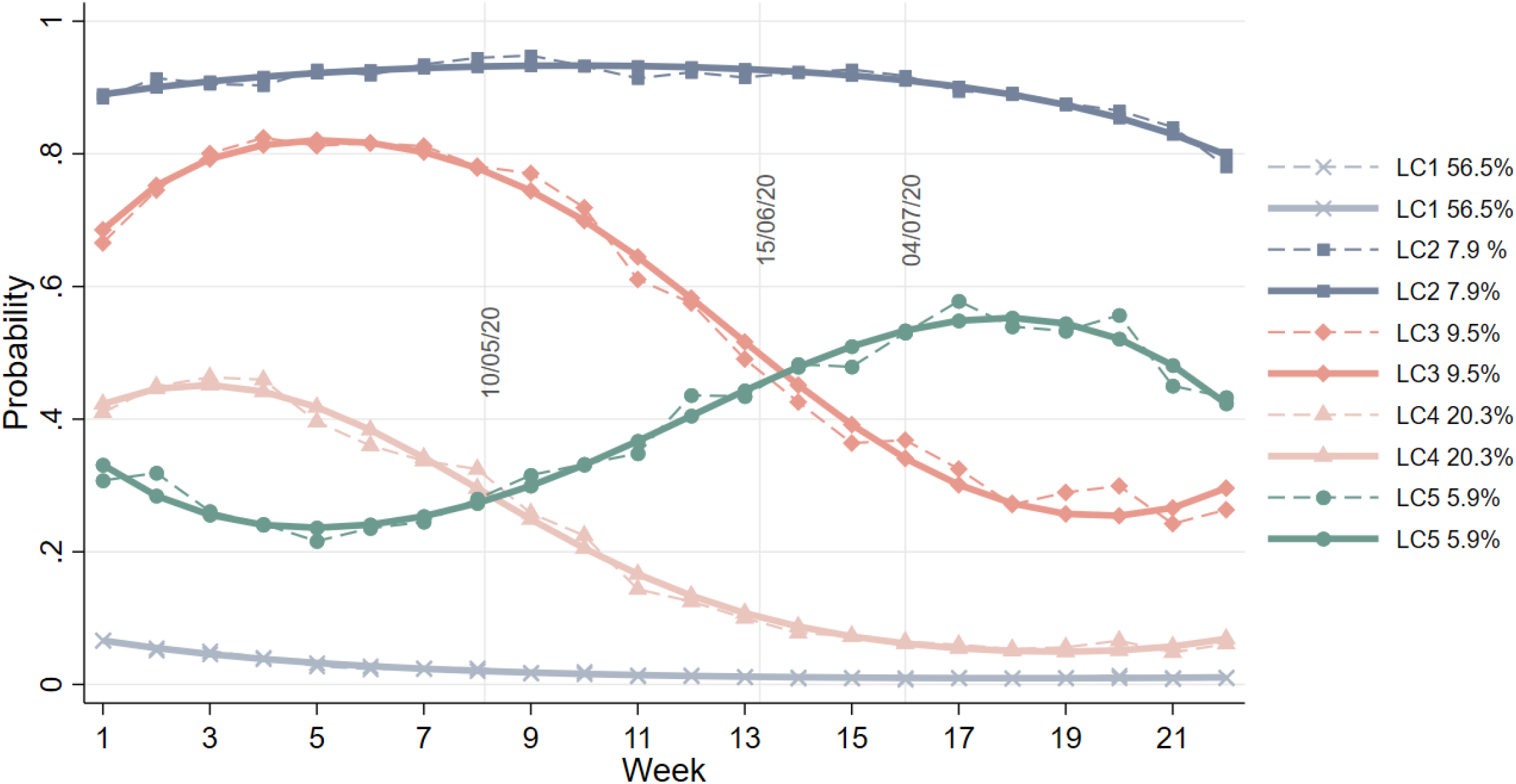
Observed (dashed lines) and estimated (solid lines) probability of arts engagement over time by latent class

### Latent trajectory classes

To determine the optimal number of latent trajectory classes, we compared across unconditional GMMs with different numbers of classes. Although the BIC and ABIC continued to decrease with each additional class added to the model, the ALMR-LR test of the six-class GMM did not reject the five-class model (Table S1). Therefore, the five-class GMM model was chosen. It had an adequate quality of class membership classification (entropy=0.82). Figure 2 shows the estimated probability of arts engagement longitudinally in each latent class (LC).

The first and largest latent class (LC1; “disengaged”; 56.5% of the sample) had a very low probability of home-based arts engagement, with little change observed over the 22-week period. In contrast, LC2 (“highly engaged”; 7.9%) included people with consistently high probabilities of arts engagement throughout the study period. The last three classes (LC3-5) were dynamic, showing substantial changes during follow-up. LC3 (“highly engaged decreasing”; 9.5%) showed an increase in the probability of home-based arts engagement in the first few weeks of lockdown, which was followed by a rapid decline as lockdown measures were eased. LC4 (“moderately engaged decreasing; 20.3%) started from a moderate probability of engaging during lockdown, which then declined sharply as lockdown measures eased, before stabilising at a very low level of engagement. Finally, LC5 (“moderately engaged increasing”; 5.9%) was the only class that showed an overall increase in arts engagement over time. In LC5, probability of arts engagement decreased slightly in the first few weeks of lockdown, steadily increased as lockdown eased, and then decreased again in the last few weeks of the study period.

### Factors associated with latent trajectory classes

We fitted a conditional GMM to examine how individual characteristics were related to the latent classes of longitudinal changes in arts engagement (Table 2). Using LC1 (“disengaged”) as the reference class, the odds of being consistently highly engaged (LC2) for women were more than threefold that for men. Compared with young adults (under 30), people aged 60+ had higher odds of being in LC2. People with a degree or above had higher odds of being in LC2 than those with GCSEs or below. Compared with people who were employed, those who lost their job during the follow-up had higher odds of being in LC2. Those who were already unemployed or economically inactive at the start of lockdown had even higher odds of being in LC2. Compared with people living alone, those living with children had higher odds of being in LC2. Additionally, people with a disability or mental health diagnosis were more likely to be in LC2 than LC1. Finally, people who reported changes in their arts engagement frequency compared to before the pandemic were more likely to be in LC2 than individuals who maintained similar engagement levels.

**Table 2.** Results from the Growth mixture model with predictors of latent classes (LC) (LC1, the disengaged, as the reference, N=29,147)

|  | Highly engaged<br>LC2 (vs. LC1) |  | Highly engaged<br>decreasing<br>LC3 (vs. LC1) |  | moderately engaged<br>decreasing<br>LC4 (vs. LC1) |  | Moderately engaged<br>increasing<br>LC5 (vs. LC1) |  |
| --- | --- | --- | --- | --- | --- | --- | --- | --- |
|  | OR | 95% CI | OR | 95% CI | OR | 95% CI | OR | 95% CI |
| Woman (vs man) | **3.09** | **[2.37-4.03]** | **3.46** | **[2.54-4.71]** | **2.36** | **[1.98-2.80]** | **2.57** | **[1.91-3.47]** |
| Ethnic minority (vs white) | 0.92 | [0.56-1.51] | 1.48 | [0.92-2.36] | 1.10 | [0.77-1.58] | 1.24 | [0.75-2.07] |
| Age 30-45 (vs. 18-29) | 1.02 | [0.66-1.58] | **0.67** | **[0.46-0.98]** | **0.67** | **[0.49-0.91]** | 0.74 | [0.47-1.16] |
| Age 46-59 (vs. 18-29) | 1.29 | [0.85-1.95] | 0.73 | [0.47-1.11] | **0.59** | **[0.43-0.80]** | **0.65** | **[0.43-0.99]** |
| Age 60+ (vs. 18-29) | **1.87** | **[1.20-2.91]** | 0.99 | [0.65-1.52] | 0.79 | [0.58-1.08] | 0.69 | [0.44-1.09] |
| Education medium (vs. low) | 1.14 | [0.87-1.49] | **1.61** | **[1.18-2.19]** | 1.07 | [0.85-1.35] | 1.13 | [0.78-1.63] |
| Education high (vs. low) | **1.33** | **[1.01-1.75]** | **1.56** | **[1.17-2.10]** | 1.15 | [0.92-1.43] | **1.45** | **[1.03-2.04]** |
| Household income 16-29k (vs. <16k) | 1.11 | [0.84-1.46] | 0.87 | [0.63-1.19] | 1.01 | [0.79-1.28] | 0.80 | [0.56-1.14] |
| Household income 30-59k (vs. <16k) | 0.75 | [0.55-1.02] | 0.79 | [0.56-1.11] | 0.98 | [0.75-1.26] | **0.46** | **[0.31-0.68]** |
| Household income 60-89k (vs. <16k) | 0.86 | [0.54-1.36] | **0.60** | **[0.38-0.95]** | 0.95 | [0.67-1.36] | **0.48** | **[0.29-0.79]** |
| Household income ≥90k (vs. <16k) | 0.67 | [0.41-1.10] | **0.53** | **[0.32-0.89]** | **0.61** | **[0.42-0.89]** | **0.45** | **[0.22-0.93]** |
| Employed to unemployed (vs. employed) | **1.51** | **[1.10-2.08]** | **1.89** | **[1.42-2.51]** | **1.54** | **[1.19-1.99]** | 1.16 | [0.76-1.78] |
| Unemployed/inactive (vs. employed) | **2.18** | **[1.69-2.80]** | **1.62** | **[1.22-2.15]** | 1.09 | [0.88-1.36] | 1.39 | [1.00-1.92] |
| Living with others, but no children (vs alone) | 1.20 | [0.96-1.50] | **1.39** | **[1.08-1.79]** | 1.11 | [0.91-1.35] | **1.38** | **[1.01-1.90]** |
| Living with others, including children (vs. alone) | **1.70** | **[1.19-2.43]** | **2.38** | **[1.76-3.21]** | **2.19** | **[1.69-2.83]** | 1.56 | [0.93-2.61] |
| Large town (vs. city) | 0.79 | [0.58-1.06] | 0.88 | [0.63-1.23] | 0.94 | [0.76-1.17] | 0.66 | [0.43-1.00] |
| Small town (vs. city) | 0.86 | [0.67-1.10] | 1.30 | [0.89-1.89] | 1.22 | [0.96-1.56] | 0.89 | [0.61-1.31] |
| Rural (vs. city) | 0.82 | [0.62-1.07] | 1.06 | [0.80-1.40] | 1.22 | [0.99-1.51] | 0.89 | [0.65-1.22] |
| Disability (vs. no disability) | **1.78** | **[1.30-2.44]** | 1.37 | [0.81-2.29] | **1.84** | **[1.40-2.42]** | 1.13 | [0.71-1.82] |
| Mental health diagnosis (vs. no diagnosis) | **1.43** | **[1.10-1.86]** | **1.68** | **[1.27-2.22]** | 1.17 | [0.95-1.45] | 1.38 | [0.98-1.93] |
| Less arts engagement than usual (vs. the same) | **1.38** | **[1.02-1.87]** | **1.60** | **[1.16-2.19]** | **1.58** | **[1.28-1.94]** | **2.05** | **[1.46-2.87]** |
| More arts engagement than usual (vs. the same) | **7.39** | **[5.88-9.30]** | **6.18** | **[4.87-7.84]** | **3.34** | **[2.75-4.05]** | **3.66** | **[2.40-5.57]** |

Note: 95% CI not including 1 in bold text

Relative to LC1 (“disengaged”), women were more likely to be in LC3 (“highly engaged decreasing”) and LC4 (“moderately engaged decreasing”). People aged 30-45 had lower odds of being in LC3 or LC4 compared to young adults, and those aged 46-59 had lower odds of being in LC4. There was no difference between young (under 30) and older adults (60+). People with a higher level of education had higher odds of being in LC3. Membership of LC3 and LC4 compared to LC1 was also related to household income. People with a household income of ≥£60,000 were less likely to be in LC3 and those with a household income of ≥£90,000 were less likely to be in LC4. Compared to the employed, people who lost their job during the pandemic had higher odds of being in LC3 and those who were unemployed or economically inactive at the start of lockdown had higher odds of being in LC3. Compared to people living alone, those who lived with other adults had higher odds of being in LC3, whereas people living with children were more likely to be in LC3 or LC4. People with a disability had higher odds of being in LC4. People with mental health diagnoses had higher odds of being in LC3. Finally, people whose arts engagement frequency changed compared to before the pandemic were more likely to be in LC3 or LC4 than individuals who maintained similar engagement levels.

Compared to LC1 (“disengaged”), women were more likely to be in LC5 (“moderately engaged increasing”). Adults aged 46-59 had lower odds of being in LC5 than young adults. People with a degree or above had higher odds of being in LC5 than those with the lowest education levels. People with a household income of ≥£30,000 were less likely to be in LC5 than those with a household income under £16,000. Compared with people living alone, those who lived with other adults had higher odds of being in LC5. Finally, people whose arts engagement frequency changed compared to before the pandemic were more likely to be in LC5 than individuals who maintained similar engagement levels.

Next, in sensitivity analyses, we altered the reference categories (Table S2). When comparing the two classes with a high probability of arts engagement at the start (LC3 “highly engaged decreasing” vs LC2 “highly engaged”), the only predictor of class membership was age. People aged 46 and over were less likely to be in LC3, indicating that they were less likely to reduce their arts engagement than young adults. Comparing the classes that started with a moderate level of arts engagement (LC5 “moderately engaged increasing” vs LC4 “moderately engaged decreasing”), people with a household income of £30,000-£89,000 were less likely to be in LC5, indicating that they were less likely to increase their engagement than individuals with the lowest income.

## Discussion

This was one of the first studies to examine patterns of longitudinal changes in home-based arts engagement during the COVID-19 pandemic, specifically exploring differences across the first UK lockdown and the easing of lockdown measures. Our analyses identified five unique classes of longitudinal changes in arts engagement. Two of these classes were stable, showing few changes as social restrictions were enforced and relaxed, including the consistently disengaged (56.5% of participants) and the consistently highly engaged (7.9%). Two classes (29.8%) showed initial increases in arts engagement during the first lockdown, followed by declines as restrictions were eased. Only one small class (5.9%) showed the opposite pattern of declines during lockdown followed by an increase as restrictions were lifted. These longitudinal changes in arts engagement are more nuanced than previously indicated by self-reported comparative measures of arts engagement [2]. We found clear changes that coincided with the easing of restrictions, suggesting that people’s motivations to engage could be directly related to policy changes.

This study further examined whether a range of factors were associated with the patterns of changes in arts engagement during the pandemic. Some factors were consistently associated with patterns of engagement, in line with previous research. Women, those with higher levels education, and those living with others were more likely to be in any group except the “disengaged”, as found both before and during the COVID-19 pandemic [2, 8, 10]. There was no evidence that ethnicity was associated with longitudinal patterns of home-based arts engagement. Pre-pandemic research has found that, when broadly defined, arts engagement differs by ethnicity [9, 10]. However, ethnic disparities may be larger in receptive cultural activities than in home-based arts activities [2, 16]. It is thus not surprising that home-based arts engagement did not differ according to ethnicity.

However, some findings were less consistent. The odds of constantly high engagement increased with age and some older age groups were less likely to have increasing or decreasing engagement than young adults. This suggests that older adults had higher and more stable levels of arts engagement. In previous studies, younger people generally engage in the arts more often [2, 17]. This inconsistency might be due to the strict lockdown measures for older adults, who were strongly advised to stay at home even after lockdown measures were relaxed for other age groups in the UK. Additional time at home might have increased opportunities and motivations for older adults to engage in arts activities to help sustain their wellbeing. Lockdown may also have caused fewer differences to normal life for retired individuals, leading to more consistent patterns of leisure engagement [18]. Younger adults might have spent more time engaging in other activities, including using social media, playing video games, and meeting others after lockdown measures were eased. Despite this, young people were more likely to have engaged in the arts than adults aged 30-59. This working age group may have faced challenges around childcare whilst working, reducing time available for leisure [19, 20].

Associations between household income and arts engagement may also have been altered by the pandemic. Pre-pandemic studies have generally demonstrated more arts engagement with increasing income [16, 21]. However, as in two recent studies [2, 17], we found that individuals with a lower household income were less likely to be in the “disengaged” group. People with lower income were also more likely to have increased arts engagement throughout the first 22 weeks of the pandemic. This may be because lower-paying jobs were more severely disrupted by the pandemic, with individuals in these roles likely to be working fewer hours [22], leaving more free time for arts engagement. However, those who were employed for the whole period were more likely to be consistently disengaged, so any form of job could still reduce time available to engage in arts activities.

People living with others were also more likely to engage in the arts. In particular, those living with children maintained high levels of engagement, although these individuals were also likely to start with high levels of engagement that declined over time. These findings are supported by the increased sales in arts and crafts supplies when schools and recreational facilities were closed [1, 7]. Arts activities might have offered new opportunities for parents to engage with their children, as well as preventing boredom at home [7]. Some of these activities were publicly visible, such as the proliferation of rainbow drawings amongst families in the UK to support frontline health professionals and key workers and to spread hope [23, 24]. It is possible that individuals living with children had decreasing levels of arts engagement throughout the pandemic due to burnout and difficulties sustaining a balance between work, childcare, schooling, and other responsibilities [19, 20]. Whilst previous studies have shown that people living in remote areas are more likely to engage in the arts [2, 25], we found no associations between living area and longitudinal patterns of arts engagement. This suggests that changes over time might not vary by level of urbanicity.

Finally, people with a disability or mental health diagnosis were more likely to be highly engaged in arts activities throughout the first 22 weeks of the pandemic. In contrast, pre-pandemic studies suggest that people with physical health conditions, lower wellbeing, and those who are less happy have lower engagement levels [10, 26, 27]. This could be due to the transition of the cultural sector to providing more opportunities for engagement online, reaching wider audiences, reducing barriers to accessing the arts, and creating new opportunities to participate, especially for people who have traditionally engaged less in the arts [2]. In addition to this greater accessibility, people with a health condition might have used arts more frequently to help manage their emotions during the pandemic [2, 28].

This study has a number of strengths including its large sample size, repeated weekly follow-up of the same participants over 22 weeks since the first UK lockdown, and robust statistical approaches. Although the UCL COVID-19 Social Study did not use a random sample, it does have a large sample size with wide heterogeneity, including good stratification across all major socio-demographic groups. In addition, analyses were weighted using population estimates of core demographics. The weighted data aligned well with national population statistics and another large nationally representative social survey [29]. Despite all efforts to make our sample inclusive and representative of the adult population in the UK, we cannot rule out the possibility of potential biases due to omitting other demographic factors that could be associated with survey participation in the weighting process. Further, our arts engagement measure focused on one weekday which might obscure possible patterns of arts engagement during weekends, especially for those who are employed. Finally, this study only investigated home-based engagement, as opportunities for arts engagement outside of the home were largely suspended. Future studies could extend our analyses by including other types of arts engagement activities and by extending the follow-up period to explore longer term patterns of arts engagement.

## Conclusions

Overall, this study provides evidence for heterogeneity in longitudinal changes in home-based arts engagement during the COVID-19 pandemic, showing five unique patterns. Only a small proportion of participants were consistently engaged in home-based arts activities. Instead, over half of the sample were consistently disengaged, and nearly a third had good levels of engagement during the first lockdown that declined as soon as lockdown eased and other activities within society resumed. Patterns of engagement could be related to changes in social restrictions, with individuals drawing on the arts when they needed them the most. Some characteristics of the audience for home-based arts activities, such as gender and education, were consistent with usual audiences for such activities. However, other groups who are usually less likely to engage in the arts, such as people with mental health conditions, engaged more during the pandemic. This is encouraging as it suggests that those who needed the arts most did indeed engage more. It also indicates audience diversification, with potential implications for the future of the cultural sector. If audiences who have traditionally been harder to reach can be engaged in times of crisis, it may be possible to encourage greater participation from them beyond the pandemic. However, as the majority of participants reverted to lower levels of engagement when restrictions eased, the effects of lockdown on arts engagement may be largely transient. Overall, these results show the importance of promoting arts engagement during pandemics as part of public health campaigns, especially when social restrictions are introduced. The engagement patterns identified suggest that even groups less likely to engage in usual circumstances have increased odds of engaging in the arts during a pandemic. Given the critical role of the arts in coping strategies, this has important ramifications for supporting public mental health. However, if the cultural sector wants to sustain changes in audiences brought about by the COVID-19 pandemic, more work is needed to re-engage those groups who have since reverted to lower levels of engagement.

## Data Availability

Anonymous data will be made publicly available following the end of the pandemic.

## Declarations

### Ethics

The study was approved by the UCL Research Ethics Committee [12467/005] and all participants gave informed consent.

### Data availability

Anonymous data will be made publicly available following the end of the pandemic.

### PPI

The research questions in the UCL COVID-19 Social Study built on patient and public involvement as part of the UKRI MARCH Mental Health Research Network, which focuses on social, cultural and community engagement and mental health. This highlighted priority research questions and measures for this study. Patients and the public were additionally involved in the recruitment of participants to the study and are actively involved in plans for the dissemination of findings from the study.

### Author contributions

FB and DF conceived and designed the study. FB analysed the data. HWM, FB and JB wrote the first draft. All authors provided critical revisions. All authors read and approved the submitted manuscript.

## References

1. Guardian (2020) Hobbycraft reports 200% boom in online sales since start of pandemic | Retail industry | The Guardian.

2. Mak HW, Fluharty M, Fancourt D Predictors and impact of arts engagement during the COVID-19 pandemic: analyses of data from 19,384 adults in the COVID-19 Social Study. https://doi.org/10.31234/OSF.IO/RCKP5

3. Fancourt D, Aughterson H, Finn S, Walker E, Steptoe A (2021) How leisure activities affect health: a narrative review and multi-level theoretical framework of mechanisms of action. The Lancet Psychiatry. https://doi.org/10.1016/S2215-0366(20)30384-9

4. Fancourt D, Finn S (2019) What is the evidence on the role of the arts in improving health and well-being? A scoping review. Copenhagen

5. Bu F, Steptoe A, Mak HW, Fancourt D (2020) Time-use and mental health during the COVID-19 pandemic: A panel analysis of 55,204 adults followed across 11 weeks of lockdown in the UK. medRxiv 2020.08.18.20177345

6. Cabedo-Mas A, Arriaga-Sanz C, Moliner-Miravet L (2021) Uses and Perceptions of Music in Times of COVID-19: A Spanish Population Survey. Front Psychol. https://doi.org/10.3389/fpsyg.2020.606180

7. Choi M, Tessler H, Kao G (2020) Arts and crafts as an educational strategy and coping mechanism for Republic of Korea and United States parents during the COVID-19 pandemic. Int Rev Educ 66:715–735

8. Devine P, Dowds L (2013) Understanding society: culture, arts and leisure in the UK regions Final Report April 2013.

9. Mak HW, Coulter R, Fancourt D (2020) Patterns of social inequality in arts and cultural participation: findings from a nationally-representative sample of adults living in the UK. WHO Public Heal Panor 6:55–68

10. Parkinson A, Buttrick J, Wallis A (2014) Equality and diversity within the arts and cultural sector in England: evidence and literature review final report.

11. Risner D (2014) Bullying victimisation and social support of adolescent male dance students: an analysis of findings. https://doi.org/10.1080/14647893.2014.891847

12. Government U (2020) £1.57 billion investment to protect Britain’s world-class cultural, arts and heritage institutions. In: UK Gov.

13. Seymour G, Malapit HJ, Quisumbing AR (2017) Measuring Time Use in Development Settings.

14. Muthén B, Asparouhov T (2008) Growth mixture modeling: Analysis with non-Gaussian random effects. In: Fitzmaurice G, Davidian M, Verbeke G, Molenberghs G (eds) Longitud. data Anal. CRC Press, Boca Raton, pp 143–165

15. Office for National Statistics Population estimates for the UK, England and Wales, Scotland and Northern Ireland. In: 2019.

16. Bone J, Bu F, Fluharty M, Paul E, Sonke J, Fancourt D (2021) Who engages in the arts in the United States? A comparison of three types of engagement using data from the General Social Survey. SocArXiv. https://doi.org/10.31235/OSF.IO/5NYQ3

17. DCMS (2020) Taking Part Web Panel COVID-19 Report.

18. McKinlay A, Fancourt D, Burton A (2020) “It makes you realise your own mortality.” A qualitative study on mental health of older adults in the UK during COVID-19. medRxiv 2020.12.15.20248238

19. Cheng Z, Mendolia S, Paloyo AR, Savage DA, Tani M (2021) Working parents, financial insecurity, and childcare: mental health in the time of COVID-19 in the UK. Rev Econ Househ 19:123–144

20. May T, Aughterson H, Fancourt D, Burton A ‘Stressed, uncomfortable, vulnerable, neglected’: a qualitative study of the psychological and social impact of the COVID-19 pandemic on UK frontline keyworkers. https://doi.org/10.31235/OSF.IO/DN43C

21. Fancourt D, Steptoe A (2019) Cultural engagement and mental health: Does socio-economic status explain the association? Soc Sci Med 236:112425

22. Wilson T, Buzzeo J (2021) Laid low and insecure workers.

23. BBC News (2020) Coronavirus: Rainbow pictures springing up across the country. BBC News

24. BBC News (2020) Coronavirus: Rainbow portraits thank the NHS. BBC News

25. Mak HW, Coulter R, Fancourt D (2020) Does arts and cultural engagement vary geographically? Evidence from the UK household longitudinal study. Public Health 185:119–126

26. Fancourt D, Baxter L (2020) Differential participation in community cultural activities amongst those with poor mental health: Analyses of the UK Taking Part Survey. Soc Sci Med 261:113221

27. Steptoe A, Fancourt D (2019) Leading a meaningful life at older ages and its relationship with social engagement, prosperity, health, biology, and time use. Proc Natl Acad Sci U S A 116:1207–1212

28. Fancourt D, Garnett C, Spiro N, West R, Müllensiefen D (2019) How do artistic creative activities regulate our emotions? Validation of the Emotion Regulation Strategies for Artistic Creative Activities Scale (ERS-ACA). PLoS One 14:1–22

29. Bu F, Steptoe A, Fancourt D (2020) Who is lonely in lockdown? Cross-cohort analyses of predictors of loneliness before and during the COVID-19 pandemic. Public Health 186:31–34

