## Supplement for "Longitudinal changes in home-based arts engagement during and following the first national lockdown due to the COVID-19 pandemic in the United Kingdom"

### Supplementary Material

#### UK Lockdown Timeline

The period between 21st March and 21st August 2020 overlapped with the first national lockdown and gradual easing of restrictions in the UK Government's response to the COVID-19 pandemic. On 23rd March, the first lockdown commenced in the UK, with people being ordered to 'stay at home'. On 10th May, it was announced that strict lockdown was being eased in England, with unlimited outdoor exercise permitted, more movement around the country permitted, and some return to work allowed. Easing of restrictions in other devolved countries are as follows: Northern Ireland on 18th May, Scotland on 29th May and Wales on 1st June. On 1st June, people from different household in England were allowed to meet outdoors under the rule of six, and schools started to reopen in phases. On 15th June, non-essential retail was reopened in England. On 4th July, further public amenities were reopened in England, and individuals from two households could meet socially indoors. On 14th August, restrictions were eased further in England, including reopening indoor theatres and bowling alleys.

#### Supplementary Tables

**Table S1.** Model fit indices for different model specifications.

| Model specification | BIC | ABIC | LMR-LR | ALMR-LR | Entropy |
|---|---|---|---|---|---|
| 1-class GMM | 443,746 | 443,733 | NA | NA | NA |
| 2-class GMM | 313,898 | 313,870 | <0.001 | <0.001 | 0.941 |
| 3-class GMM | 294,732 | 294,687 | <0.001 | <0.001 | 0.886 |
| 4-class GMM | 290,612 | 290,551 | <0.001 | <0.001 | 0.822 |
| **5-class GMM** | **287,220** | **287,144** | **0.020** | **0.023** | **0.819** |
| 6-class GMM | 286,115 | 286,023 | 0.204 | 0.208 | 0.757 |

**Table S2.** Results from the growth mixture model including predictors of latent classes (LC) using alternative reference classes (N=29,147).

| | LC3 (vs. LC2) | | LC3 (vs. LC4) | | LC5 (vs. LC4) | |
|---|---|---|---|---|---|---|
| | OR | 95% CI | OR | 95% CI | OR | 95% CI |
| Woman (vs man) | 1.12 | [0.75-1.68] | **1.47** | **[1.04-2.08]** | 1.09 | [0.77-1.54] |
| Ethnic minority (vs white) | 1.60 | [0.82-3.15] | 1.34 | [0.80-2.24] | 1.13 | [0.63-2.02] |
| Age 30-45 (vs. 18-29) | 0.66 | [0.38-1.15] | 1.01 | [0.66-1.54] | 1.11 | [0.66-1.86] |
| Age 46-59 (vs. 18-29) | **0.56** | **[0.32-0.99]** | 1.23 | [0.78-1.96] | 1.10 | [0.69-1.78] |
| Age 60+ (vs. 18-29) | **0.53** | **[0.30-0.96]** | 1.25 | [0.79-1.99] | 0.87 | [0.53-1.44] |
| Education medium (vs. low) | 1.41 | [0.94-2.10] | **1.50** | **[1.06-2.12]** | 1.05 | [0.68-1.62] |
| Education high (vs. low) | 1.18 | [0.80-1.74] | 1.36 | [0.99-1.88] | 1.26 | [0.86-1.85] |
| Household income 16-29k (vs. <16k) | 0.78 | [0.52-1.18] | 0.86 | [0.60-1.23] | 0.79 | [0.53-1.19] |
| Household income 30-59k (vs. <16k) | 1.05 | [0.67-1.66] | 0.81 | [0.55-1.19] | **0.47** | **[0.31-0.72]** |
| Household income 60-89k (vs. <16k) | 0.70 | [0.36-1.36] | 0.64 | [0.37-1.10] | **0.50** | **[0.28-0.89]** |
| Household income ≥90k (vs. <16k) | 0.80 | [0.40-1.59] | 0.87 | [0.50-1.53] | 0.74 | [0.35-1.56] |
| Employed to unemployed (vs. employed) | 1.25 | [0.85-1.84] | 1.23 | [0.89-1.69] | 0.76 | [0.47-1.21] |
| Unemployed/inactive (vs. employed) | 0.74 | [0.52-1.07] | **1.48** | **[1.08-2.03]** | 1.27 | [0.88-1.84] |
| Living with others, but no children (vs alone) | 1.16 | [0.84-1.60] | 1.26 | [0.94-1.68] | 1.25 | [0.87-1.78] |
| Living with others, including children (vs. alone) | 1.40 | [0.90-2.17] | 1.09 | [0.77-1.54] | 0.71 | [0.40-1.26] |
| Large town (vs. city) | 1.12 | [0.72-1.76] | 0.94 | [0.65-1.34] | 0.70 | [0.44-1.10] |
| Small town (vs. city) | 1.51 | [0.98-2.30] | 1.06 | [0.69-1.63] | 0.73 | [0.46-1.16] |
| Rural (vs. city) | 1.29 | [0.90-1.85] | 0.87 | [0.63-1.19] | 0.73 | [0.51-1.04] |
| Disability (vs. no disability) | 0.77 | [0.44-1.33] | 0.74 | [0.44-1.25] | 0.62 | [0.36-1.05] |
| Mental health diagnosis (vs. no diagnosis) | 1.18 | [0.82-1.70] | **1.43** | **[1.06-1.94]** | 1.17 | [0.81-1.71] |
| Less arts engagement than usual (vs. the same) | 1.15 | [0.75-1.78] | 1.01 | [0.72-1.43] | 1.30 | [0.90-1.88] |
| More arts engagement than usual (vs. the same) | 0.84 | [0.61-1.15] | **1.85** | **[1.43-2.40]** | 1.10 | [0.68-1.77] |

Note: 95% CI not including 1 in bold text
